# Urinary Clusterin is a biomarker of renal epithelial senescence and predicts human kidney disease progression

**DOI:** 10.1101/2024.03.14.24303997

**Authors:** David Baird, Maximillian Reck, Ross Campbell, Marie-Helena Docherty, Matthieu Vermeren, Andy Nam, Wei Yang, Nathan Schurman, Claire Williams, Jamie P. Traynor, Patrick B. Mark, Katie Mylonas, Jeremy Hughes, Laura Denby, Bryan Conway, David A Ferenbach

## Abstract

Cellular senescence drives organ fibrosis and ageing, and accumulating evidence supports the ability of senescence-depleting drugs to improve outcomes in experimental models of disease. The lack of non-invasive biomarkers represents a major obstacle to the design of human trials of candidate senolytics. On samples from 51 patients with chronic kidney disease (CKD), we performed liquid chromatography mass spectrometry (LC-MS) analysis of urine samples alongside immunofluorescence staining of paired kidney biopsies for p21, Ki67, and CD10+Pancytokeratin as senescence, proliferation and pan-epithelial cell markers respectively. Only Urinary Clusterin (uClusterin) correlated tightly with p21+ epithelial senescence *in vivo* (rho >0.5, p<0.001) and was upregulated in the *in vitro* SASP atlas. This was validated in a second cohort of matched urine and kidney samples from n=53 participants, with uClusterin predicting levels of senescence after adjusting for renal function, age and albuminuria. In spatial transcriptomic data from n=13 CKD patients, Clusterin colocalised with senescence marker CDKN1A. In a larger cohort of n=322 participants, elevated levels of uClusterin predicted CKD progression (defined as reaching ESKD or >40% reduction in renal function) after adjusting for baseline eGFR, albuminuria, age, systolic blood pressure (SBP) and sex. uClusterin levels represents a surrogate for histological quantification of p21+Ki67- senescent renal epithelia and predicts outcomes in human kidney disease independent of existing clinical risk factors.

---

Cellular senescence drives organ fibrosis and ageing, and accumulating evidence supports the ability of senescence-depleting drugs (‘senolytics’) to improve outcomes in experimental models of disease(1). With no non-invasive biomarkers available, quantifying senescence in human solid organs requires tissue biopsy to identify cells expressing cyclin dependent kinase inhibitors p21 and/or p16^INK4A^ in the absence of proliferation markers. This represents a major obstacle to the design of human trials of candidate senolytics and assessment of treatment responses.

In human chronic kidney disease (CKD), a condition affecting over 850 million people worldwide, p21 protein expression is upregulated in sub-populations of renal epithelia. Levels of *CDKN1A* (encoding p21) transcript correlate with worsened kidney function, (2). The Kidney Precision Medicine Project dataset demonstrated upregulated *CDKN1A* (p21) expression in ‘late-adaptive’ proximal tubular (PT) epithelia in human CKD (3). *In vitro,* irradiated, Ki67 negative human PT cells expressing p21 upregulated multiple senescence-associated transcripts (2). *In vivo*, our recent multi-omic atlas of human kidney disease confirmed that CDKN1A (p21) expression was upregulated in a proximal tubular subset expressing key DEGs of KPMP ‘late-adaptive’ PTs’ alongside multiple senescence-associated DEGs(4).

We addressed the hypothesis that senescent renal epithelia could be quantified non-invasively by detecting their selectively secreted proteins in human urine samples

To identify urinary proteins correlating with tubular cell senescence, we first performed high-performance liquid chromatography with tandem mass spectrometry (LC-MS) analysis of 51 urine samples from the <u>N</u>on-inva<u>s</u>ive bi<u>o</u>markers of renal disease (seNSOR) CKD biobank (‘discovery’ cohort, Fig A, Fig S1A, Supplemental materials and methods, Table S1A), alongside staining of paired kidney biopsies for p21, Ki67, and CD10+Pancytokeratin as senescence, proliferation and pan-epithelial cell markers respectively (Fig B, Fig S1B). 331 distinct proteins were detected by LC-MS (Table S2), of which 81 had a positive correlation with proportion of p21**^+^**Ki67**^-^** senescent epithelia (adj p<0.05) with 8 having a correlation coefficient (rho)>0.5 (Fig C). Results were compared to proteins upregulated in the ‘SASP atlas’ of senescent human renal epithelia *in vitro* (*5*). Urinary Clusterin (uClusterin) alone correlated tightly with p21+ epithelial senescence *in vivo* (Fig C, rho >0.53, p<0.001) and was upregulated in the *in vitro* SASP atlas (5). p21**^+^**Ki67**^-^** senescent epithelial proportions also correlated with increasing patient age and urinary albumin:creatinine ratio (uACR) and inversely with estimated glomerular filtration rate (eGFR) (Fig S1C-E). uClusterin was quantified using ELISA in this discovery cohort and corrected for urinary concentration against urinary creatinine (Fig S1F). Multivariate analysis demonstrated that uClusterin:Creatinine levels predicted p21+Ki67- senescent renal epithelia levels after correction for eGFR, Age and uACR (Fig D).

**Figure:**
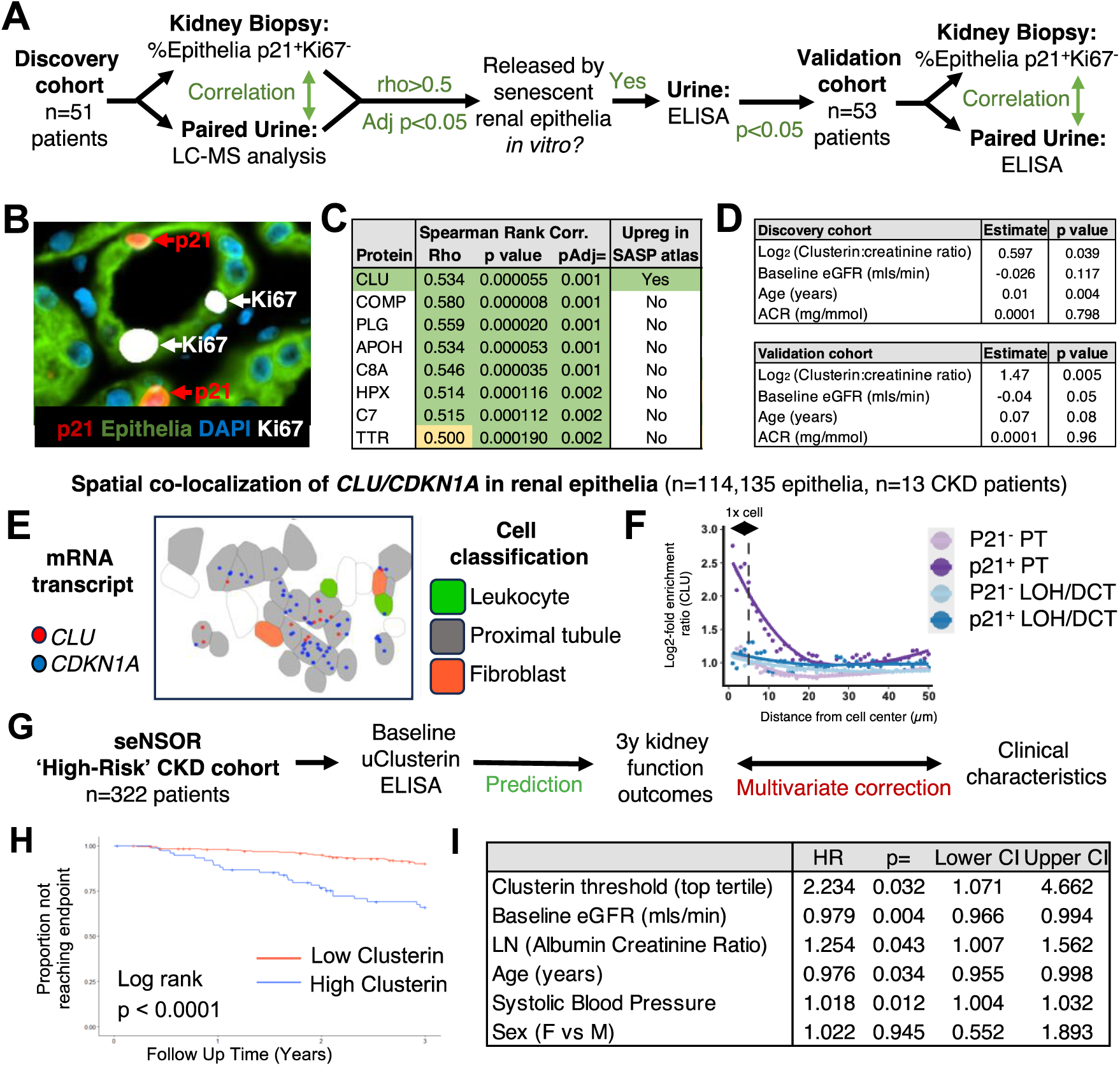
Discovery and validation of urinary Clusterin as a biomarker of renal senescence and patient outcomes. (**A**) Schema of cohort design for Discovery and Validation of urinary biomarkers of p21+Ki67- senescent epithelia. (**B**) Immunofluorescent staining of human renal biopsies. (**C**) Urinary proteins in LC-MS analysis correlating most closely with the proportion of p21+Ki67- senescent epithelial cells. (**D**) Discovery and Validation cohort linear regression analysis for predicting p21+Ki67- senescent epithelial cell proportions. (**E**) Image of CLU/CDKN1A co-localisation by spatial transcriptomic analysis. (**F**) Spatial enrichment (log2 scale) of CLU in proximity to p21^+^ and p21^-^ renal epithelial cells. (**G**) Schema of outcome studies in human CKD. (**H**) Proportion of patients with renal endpoint stratified by low (<124.5 µg/mmol) or high (>124.5 µg/mmol) Clusterin level. (**I**) Cox proportional hazards regression in outcome cohort.

Studies have shown increased Clusterin secretion from human cells undergoing senescence *in vivo* in other organs (6). To determine whether Clusterin production was selectively increased in p21**^+^**Ki67**^-^** senescent renal epithelia, sub-cellular resolution spatial transcriptomic analysis (Nanostring CosMx) was performed on kidney tissue from 13 patients with CKD. CLU transcript levels were increased within CDKN1A expressing proximal tubular epithelia on automated counting and co-localisation analysis (n=114,135 epithelia, pAdj=3.91 x 10^-8^, Figs E & F).

Given the spatial transcript co-localisation between CLU and CDKN1A and the correlation between urinary Clusterin and histological evidence of p21+ growth arrest, we confirmed our findings in a validation cohort of CKD patients from Glasgow (Fig A, Table S1A). Urinary Clusterin:creatinine ratio again correlated strongly with p21**^+^**Ki67**^-^** senescent epithelial cell proportions (rho=0.61, p<0.001, Fig S1G), with adjusted uClusterin:creatinine levels predicting histological senescence after correction for eGFR, uACR and Age (Fig D).

Next, we explored the optimal uClusterin:Creatinine ratio required to identify patients with high levels of p21**^+^**Ki67**^-^** senescent epithelia. By prioritising high specificity over high sensitivity, a threshold of 124.5 µg Clusterin/mmol Creatinine was selected. This corresponded to a sensitivity of 68% and specificity of 90% for identifying those with the highest tertile of p21**^+^**Ki67**^-^** senescent epithelia, with the AUC of the receiver operating characteristic (ROC) 81.2% (Fig S1H).

We tested whether the uClusterin:Creatinine ratio predicted a decline in renal function in 322 patients with CKD (Fig G, Fig S1A). The composite CKD progression endpoint (defined as reaching ESKD or >40% reduction in renal function from eGFR at baseline) occurred in 47 (15%) participants during 3 years follow-up.

76 participants had uClusterin:Creatinine levels >124.5 µg/mmol. These participants had a higher risk of CKD progression (log-rank p < 0.001, Fig H). In Cox Proportional Hazards analysis, a uClusterin:Creatinine level above 124.5 µg/mmol predicted CKD progression after adjusting for baseline eGFR, uACR, age, systolic blood pressure (SBP) and sex in multivariate analysis (HR 2.2, 95% C.I. 1.07-4.66, p=0.03, Fig I). Including uClusterin:Creatinine levels as continuous variables (either untransformed or log-transformed) in models was also significant and additive to biomarkers currently used in clinical practice for predicting CKD progression in multivariate analyses (Table S3).

This study demonstrates that measurement of uClusterin:Creatinine levels represents a surrogate for histological quantification of p21**^+^**Ki67**^-^** senescent renal epithelia and predicts outcomes in human kidney disease independent of and additively to existing clinical risk factors. These findings provide additional evidence connecting human renal epithelial senescence to functional decline in kidney disease. They also demonstrate the ability of non-invasive biomarkers to quantify senescent cells in solid organs and identify patients at elevated risk of progressive kidney disease for trial recruitment and targeting of senolytic therapies and monitoring of therapeutic response.

## Data Availability

All data produced in the present study are available upon reasonable request to the authors.

## Supplementary Materials and Methods

### Sex as a biological variable

Our study examined male and female human patients with CKD, and findings are reported from both sexes.

### seNSOR biobank recruitment

The seNSOR biobank comprised 635 patients recruited from renal clinics at the Royal Infirmary of Edinburgh, Edinburgh between March 2017 and March 2019. In addition, 100 patients with CKD were recruited to seNSOR from the Queen Elizabeth University Hospital, Glasgow between March 2018 and August 2019. Ethical approval was obtained from the Offices for Research Ethics Committee (REC/15/ES/0094, REC/20/ES/0061, REC 14/WS/1035 and REC 22/WS/0020) and informed patient consent and anonymisation undertaken in line with the Uniform Requirements of the International Committee of Medical Journal Editors as described previously. Participant information was collected upon enrollment with urine samples snap-frozen and stored at −80 °C.

In both centres, participant information collected upon enrolment included age, ethnicity, blood pressure and aetiology of CKD. Baseline and follow-up laboratory data obtained included serum creatinine. Glomerular filtration rate was estimated from serum creatinine using the 2009 Chronic Kidney Disease Epidemiology Collaboration (CKD-EPI) equation (1).

### Patient selection for discovery and validation cohorts

The discovery cohort of 51 participants included all those recruited in Edinburgh with matched kidney tissue and urine samples available for analysis. The validation cohort included 53 participants that were recruited in Glasgow and also had matched kidney tissue and urine samples available. All kidney biopsies were performed for clinical indications by physicians independent of research team.

### Patient selection for outcome analysis

Urine samples were available from 570 participants recruited into the seNSOR. 129 patients at low risk of CKD progression (baseline eGFR > 60 mls/min and ACR < 30mg/mmol) were excluded (Fig S1A). This matches the criterion used by other CKD biobanks (2). An additional 119 with baseline eGFR < 20 mls/min were excluded. The remaining 322 participants were included in the outcome analysis.

### LC-MS analysis

LC-MS studies were undertaken on all urine samples in the discovery cohort by Lisa Imrie and Tessa Moses from the Edinomics Team (University of Edinburgh). Samples were depleted of high abundance proteins (serum albumin and IgG) using Agilent multiple affinity removal spin (MARS) cartridges following manufacturers protocol. They were trypsin digested using S-TrapTM (Protifi) following manufacturers protocol. After speed vac drying, peptide samples were re-suspended in MS-loading buffer (0.05% v/v trifluoroacetic acid in water) and 50 pmol of MassPREP Alcohol dehydrogenase (ADH) digestion standard (Waters) was spiked into each sample (added as an external standard). They were then filtered using Millex filter before HPLC-MS analysis.

Nano- Electrospray ionization (ESI)- High-performance liquid chromatography (HPLC)- MS/MS analysis was performed using an online system of a nano-HPLC (Dionex Ultimate 3000 RSLC, Thermo-Fisher Scientific) coupled to a QExactive mass spectrometer (Thermo-Fisher Scientific) with a 300 μm x 5 mm pre-column (Acclaim Pepmap, 5 μm particle size) joined with a 75 μm x 50 cm column (EASY- Spray, 3 μm particle size). The nano-pump was run using solvent A (2%Acetonitrile in water 0.1% formic acid) and solvent B (80% acetonitrile-20% water and 0.1% formic acid) and peptides were separated using a multi-step gradient of 2–98% buffer B at a flow rate of 300 nL/min over 90 min. Progenesis (version 4 Nonlinear Dynamics, UK) was used for LC-MS label-free quantitation. Filtering was carried out so that only MS/MS peaks with a charge of 2+, 3+ or 4+ were taken into account for the total number of ‘features’ (signal at one particular retention time and m/z) and only the five most intense spectra per ‘feature’ were included. MS/MS spectra was searched using MASCOT Version 2.4 (Matrix Science Ltd) against a UniProt *H.sapiens* database with maximum missed-cut value set to 2. The following parameters were used in all searches: i) variable methionine oxidation, ii) fixed cysteine carbamidomethylation, iii) precursor mass tolerance of 10 ppm, iv) MS/MS tolerance of 0.05 Da, v) significance threshold (p) below 0.05 and vi) final peptide score of 20. Only proteins with 2 or more unique peptides were considered.

### ELISA analysis

Urine Clusterin was measured on stored urine samples using R&D Duoset ELISAs (R&D Systems, Minneapolis, MN, catalogue number DY5874) with all samples run in duplicate. Based on pilot studies most samples were analysed following 1:1000 pre-dilution, with any results outside the published working range of the assay rerun at 1:100, 1 in 10,000 or 1:100,000 dilution.

### Biochemical assays

Urinary creatinine measurements were determined using the creatininase/creatinase enzymatic method making use of a commercial kit (17654H, Sentinel Diagnostics via Alpha Laboratories Ltd., Eastleigh, UK) adapted for use on either a Cobas Fara or Mira analyser (Roche). Intra-assay precision was < 3% while inter-assay precision was CV < 5%. Urinary Clusterin was corrected for urinary creatinine throughout.

Microalbumin measurements were determined using a commercial kit (#1 0242 99 10 021, DiaSys Diagnostic Systems) adapted for use on a Cobas Mira analyser (Roche). This immunoturbidimetric assay was standardised against purified mouse albumin standards (Sigma Aldrich) with samples diluted in deionised water as appropriate. Intra-assay precision was < 5% while inter-assay precision was < 7.1%.

### Tissue biopsy staining

Immunofluorescence staining on human kidney biopsy samples was performed using a BOND III automated immunostainer (Leica Biosystems) using sequential Tyramide-coupled fluorophores. Antibody concentrations were determined by titration of single immunofluorescent stains to determine the most suitable dilution before being combined into a multiplex stain. The immunofluorescent stain comprised 4 antibodies. Pancytokeratin (CKPAN, Merck, Cat C2562) at 1:6000 dilution, and CD10 (Leica, Cat: NCL-L-CD10-270) at 1:600, both tubular epithelial markers, were added together, and the same Opal 520 fluorophore used for both. Other antibodies used in sequence targeted Ki67 (Agilent, Cat M724001-2) with an Opal 650 fluorophore and p21(Abcam, Cat ab109520) with an Opal 570 fluorophore (all secondary antibodies used at 1:500 dilution).

### Microscopy and image analysis

Images of whole slides were acquired using the Axio Scan Z1 whole slide scanner (Zeiss, Jena, Germany). Images were then analysed using QUPath (version 0.3.2). Cells were classified automatically using the algorithm below based on nuclear staining of p21 and Ki67 as follows; p21 positive/Ki67 negative, Ki67 negative/p21 positive, double positive or double negative. Only tubular epithelia that were p21 positive and Ki67 negative were classed as having a cell cycle inhibitor profile consistent with senescence and this number was expressed as a percentage of all tubular cells. In participants where >1 section from their biopsy was available, the cell counts from each section were combined before percentages were calculated.

setImageType(’FLUORESCENCE’);

resetSelection(); createAnnotationsFromPixelClassifier(“FindTissue”, 0.0, 0.0)

selectAnnotations();

runPlugin(’qupath.imagej.detect.cells.WatershedCellDetection’, ‘{“detectionImage”:

“DAPI”, “requestedPixelSizeMicrons”: 0.5, “backgroundRadiusMicrons”: 8.0,

“medianRadiusMicrons”: 0.0, “sigmaMicrons”: 1.5, “minAreaMicrons”: 10.0,

“maxAreaMicrons”: 400.0, “threshold”: 400.0, “watershedPostProcess”: true,

“cellExpansionMicrons”: 5.0, “includeNuclei”: true, “smoothBoundaries”: true,

“makeMeasurements”: true}’);

createAnnotationsFromPixelClassifier(“FindTubules”, 0.0, 0.0)

runObjectClassifier(“DB_AI_6”);

### Spatial transcriptomic analysis

Spatial transcriptomic analysis was performed on kidney tissue from 13 patients; this included 9 core biopsies from patients in the discovery cohort (n=3 with minimal change disease and n = 6 with IgA nephropathy) and an additional 4 nephrectomy specimens from patients with fibrosis due to recurrent pyelonephritis (Table S1B). CDKN1A and CLU expression in sub-cellular resolution spatial transcriptomics data (NanoString CosMx SMI) was analysed using pre-processed data downloaded in Seurat (4.4.0) format from gene expression omnibus (GSE253439). The original cell annotations were used to classify proximal tubule (PT), or loop of Henle and distal convoluted tubule (LOH-DCT) cells which express CDKN1A (independent of cell state). To avoid false positive classifications due to noise in the assay and cell segmentation errors, cells were considered CDKN1A+ when at least 2 or more CDKN1A transcripts were detected within cell segmentation boundaries.

Spatial enrichment of CLU transcripts in relation to cell centroids of a given cell type were calculated as described previously (3). Briefly, for each individual cell, a search radius of 50μm in 1μm steps from the cell centroid was defined. For each given cell type, the number detected transcripts (of a given gene) were summed and normalised by the area of the circle segment and the number of cells encountered in the search area. Simultaneously, to calculate the background signal in the general cell population, 10,000 random cells were selected and the normalised transcript counts were calculated as before. The enrichment ratio at each interval was then defined as the log2+1-fold enrichment ratio of the query cell type over the randomly selected cell population. Cell boundaries and transcripts in 2D coordinates were visualised using the Seurat ImageDimPlot function.

Differential gene expression between CDKN1A+ and CDKN1A- PT cells was assessed using the Wilcoxon signed-rank test implemented by the Seurat function FindMarkers() with default parameters.

### Statistical tests

Normality was assessed by Shapiro-Wilk test for all variables. Clinical characteristics for continuous data were expressed as mean ± standard deviation when data was normally distributed and median (interquartile range) when not normally distributed. Categorical variables were expressed as counts. When comparing two unpaired groups, a T-test was used when the data was normally distributed, and a Mann-Whitney test used if the data was not normally distributed. Categorical values were assessed using a Chi-square test.

For the LC-MS data, values were corrected for ADH and then for urinary creatinine. To determine the linear correlation between each protein detected and histological p21+Ki67- epithelial cell proportions, correlation coefficients (rho) were estimated using Spearman’s rank tests as the data was not normally distributed. Adjusted p values were calculated using the Benjamini and Hochberg False Discovery Rate (FDR) method from the list of p values, generated from the correlation between p21+Ki67- epithelial cell proportions and protein levels..

Linear regression was used to determine if levels of log2-tranformed Clusterin predicted histological p21+Ki67- epithelial cell proportions as the dependent variable in models alongside baseline eGFR, ACR and patient age.

Receiver operating characteristic (ROC) curve analysis was used to explore discrimination between those with top tertile Clusterin:Creatinine levels and determine the optimal cutoff point.

For the outcome analysis, CKD progression was defined as reaching ESKD (starting renal replacement therapy (RRT) or maintaining an eGFR <15mls/min for >90 days) or >40% reduction in renal function from eGFR at baseline (maintained for >90 days) (4) (5). Kaplan-Meier survival curves were constructed with the log-rank test used to compare curves. Univariate and multivariate analyses of outcomes using Cox proportional hazards survival models were performed. Death was treated as a censoring event. The proportional hazards assumption was tested and valid. A p value of less than 0.05 was considered significant.

All tests were performed using R version 4.1.2 or Graphpad Prism version 9. Funding: DB is supported by MR/W00089X/1, DF is supported by MR/X006735/1. seNSOR is funded by RP_042_20160304 awarded to LD. LD is supported by SF_001_20181122

---

**Supplementary Figure 1.**
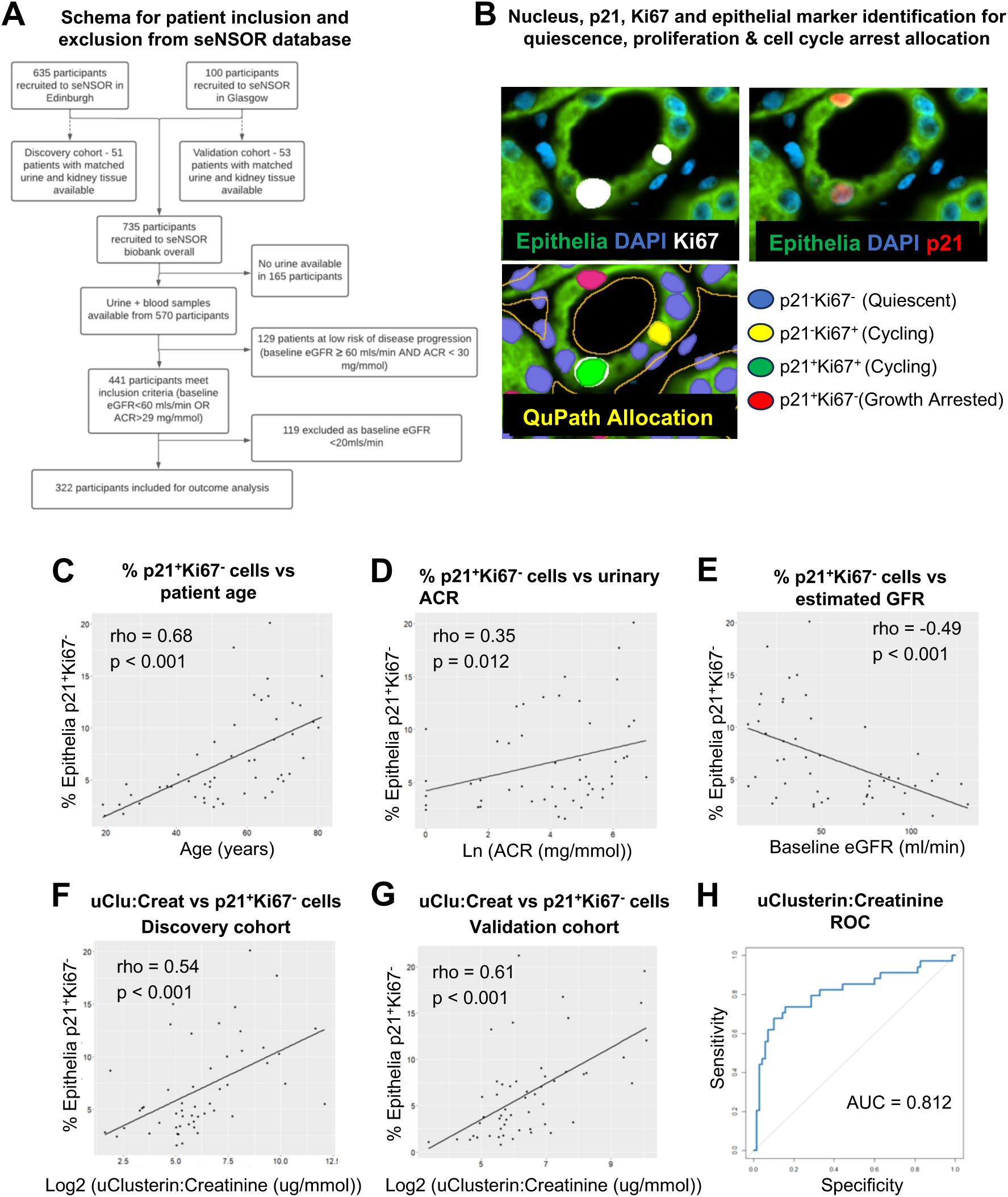
(**A**) Schema of patients included from the seNSOR biobank for the discovery, validation, and outcome analysis cohorts. (**B**) Immunofluorescent staining of human renal biopsies, same area as shown in Figure B but with selected channels as indicated and the QuPath annotations superimposed. (**C**) Correlation in the discovery cohort between the proportion of p21+Ki67- senescent epithelial cells and age. (**D**) Correlation in the discovery cohort between the proportion of p21+Ki67- senescent epithelial cells and ACR (**E**). Correlation in the discovery cohort between the proportion of p21+Ki67- senescent epithelial cells and estimated GFR. (**F**) Correlation between the proportion of p21+Ki67- senescent epithelial cells and urinary Clusterin:creatinine in the discovery cohort. (**G**) Correlation between the proportion of p21+Ki67- senescent epithelial cells and urinary Clusterin:creatinine in the validation cohort. (**H**) Receiver operating characteristic curve for urinary Clusterin:creatinine results discriminating between those in the highest tertile of p21+Ki67- senescent epithelial proportions and other participants.

**Supplemental Table S1A.** Baseline characteristics of discovery and validation cohorts.

|  | Discovery | Validation | p value for comparison |
| --- | --- | --- | --- |
| N | 51 | 53 |  |
| Age (years), median (IQR) | 56.2 (45.6, 67.4) | 54.9 (37.1, 65.3) | 0.61 |
| Male/Female, n | 32/19 | 29/24 | 0.53 |
| eGFR (ml/min), median (IQR) | 45.6 (28.8, 83.8) | 47.1 (32.6, 84.2) | 0.61 |
| SBP, mean ± S.D | 139 ± 21.8 | 139.5 ± 20.5 | 0.9 |
| DBP, mean ± S.D | 80.2 ± 12.4 | 82.5 ± 13.6 | 0.37 |
| uACR (mg/mmol), median (IQR) | 85.9 (16.4, 247.7) | 131.2 (29.2, 333.2) | 0.14 |
| Ethnicity, |  |  |  |
| White, n | 47 | 43 |  |
| Black, n | 1 | 0 |  |
| Asian, n | 1 | 3 |  |
| Not recorded, n | 2 | 7 |  |
| Diagnoses, |  |  |  |
| IgA Nephropathy | 21 | 9 |  |
| Membranous Nephropathy | 0 | 7 |  |
| Interstitial Nephritis | 6 | 8 |  |
| Minimal Change Disease | 5 | 1 |  |
| Primary FSGS | 0 | 4 |  |
| Diabetic Nephropathy | 5 | 3 |  |
| Vasculitis | 5 | 3 |  |
| Hypertensive / ischaemic nephropathy | 2 | 5 |  |
| Lupus nephritis | 2 | 3 |  |
| Other * | 5 | 10 |  |

\* Other diagnoses in discovery cohort: Henoch-Schönlein purpura (n=2), lithium toxicity, obesity glomerulopathy and myeloma, and validation cohort: mesangiocapillary glomerulonephritis (n=2), thin glomerular basement membrane (n=2), Henoch-Schönlein purpura, AA amyloid, AL amyloid, infiltration by lymphoma, CKD of uncertain aetiology and acute kidney injury.

**Supplemental Table S1B.** Baseline characteristics of outcome analysis cohorts.

|  | Outcome cohort |
| --- | --- |
| N | 322 |
| Age (years), median (IQR). | 60.5 (47.9, 69.3) |
| Male/Female, n | 192/130 |
| eGFR (ml/min/1.73m<sup>2</sup>), median (IQR) | 42.1 (29.8, 64.4) |
| SBP, median (IQR) | 135 (122, 148) |
| DBP, median (IQR) | 78 (70, 84) |
| uACR (mg/mmol), median (IQR) | 81.6 (14.5, 288.8) |
| Ethnicity, |  |
| White, n | 279 |
| Black, n | 5 |
| Asian, n | 12 |
| Not recorded, n | 26 |
| Diagnoses, |  |
| Glomerular Disease | 153 |
| Tubulointerstitial Disease | 44 |
| Diabetes Mellitus | 29 |
| Renovascular disease / Hypertension | 27 |
| Other systemic diseases affecting kidney | 7 |
| Familial / Hereditary Nephropathies | 10 |
| Miscellaneous Renal Disorders | 52 |

**Supplemental Table S2.**
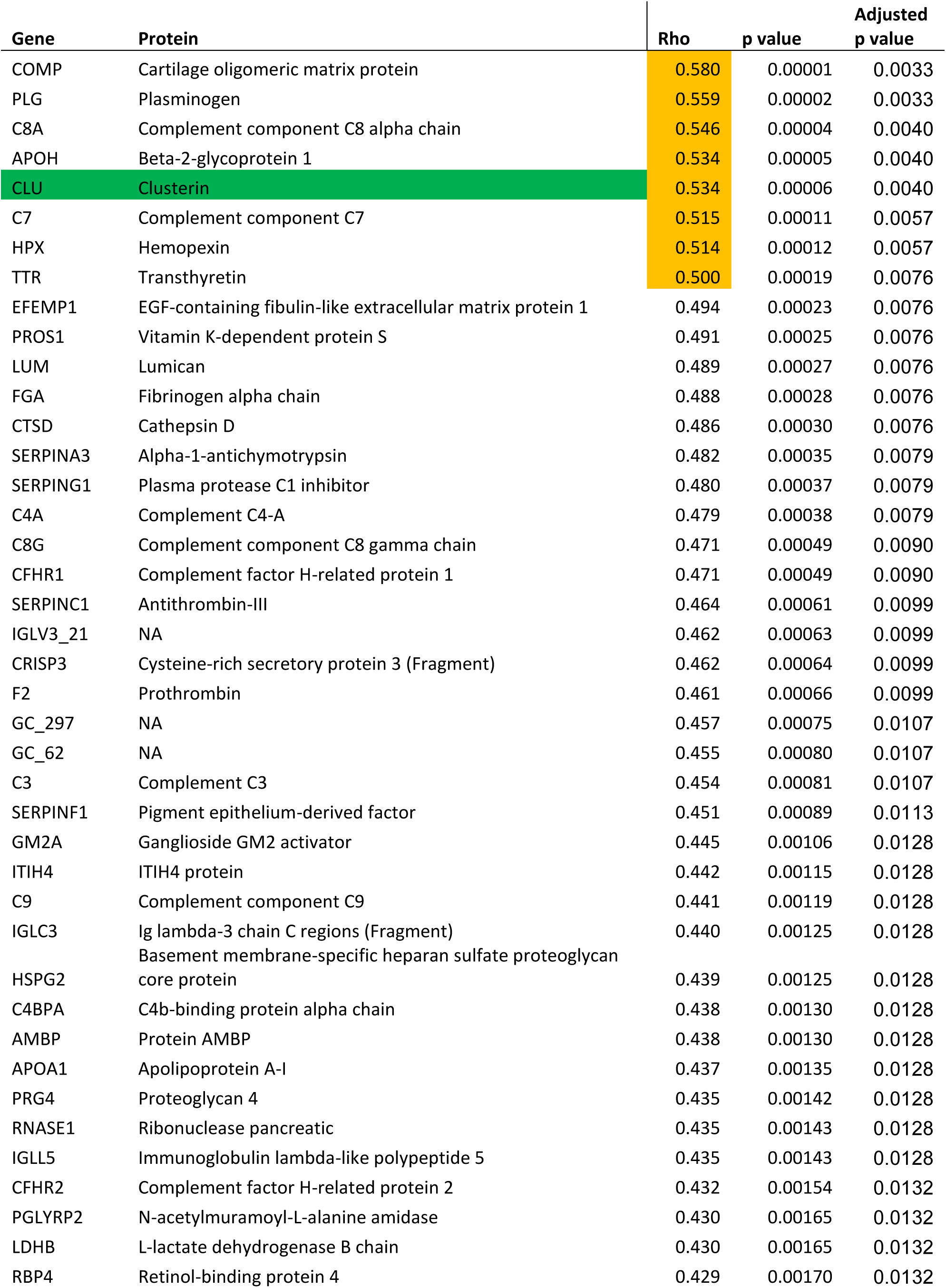
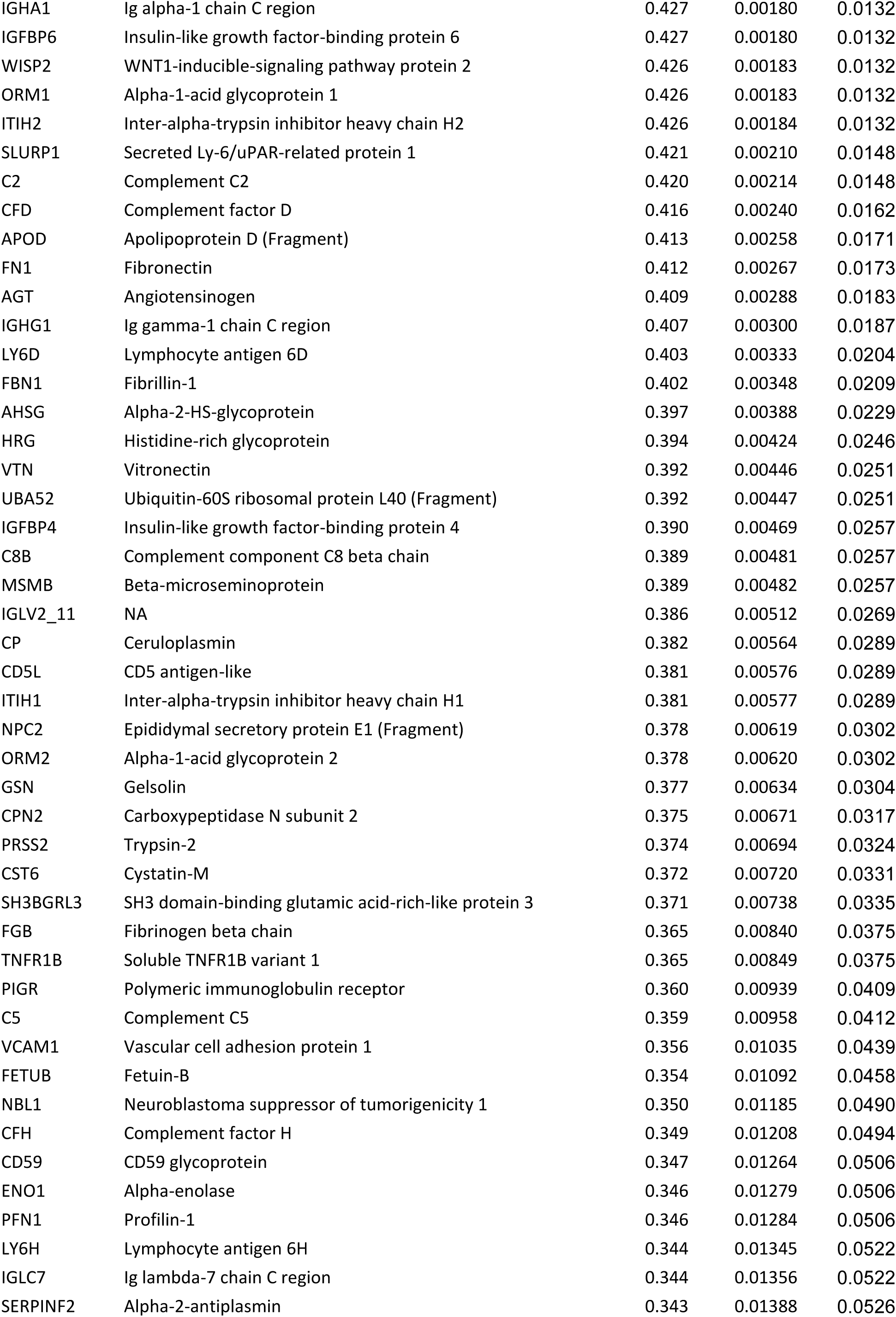
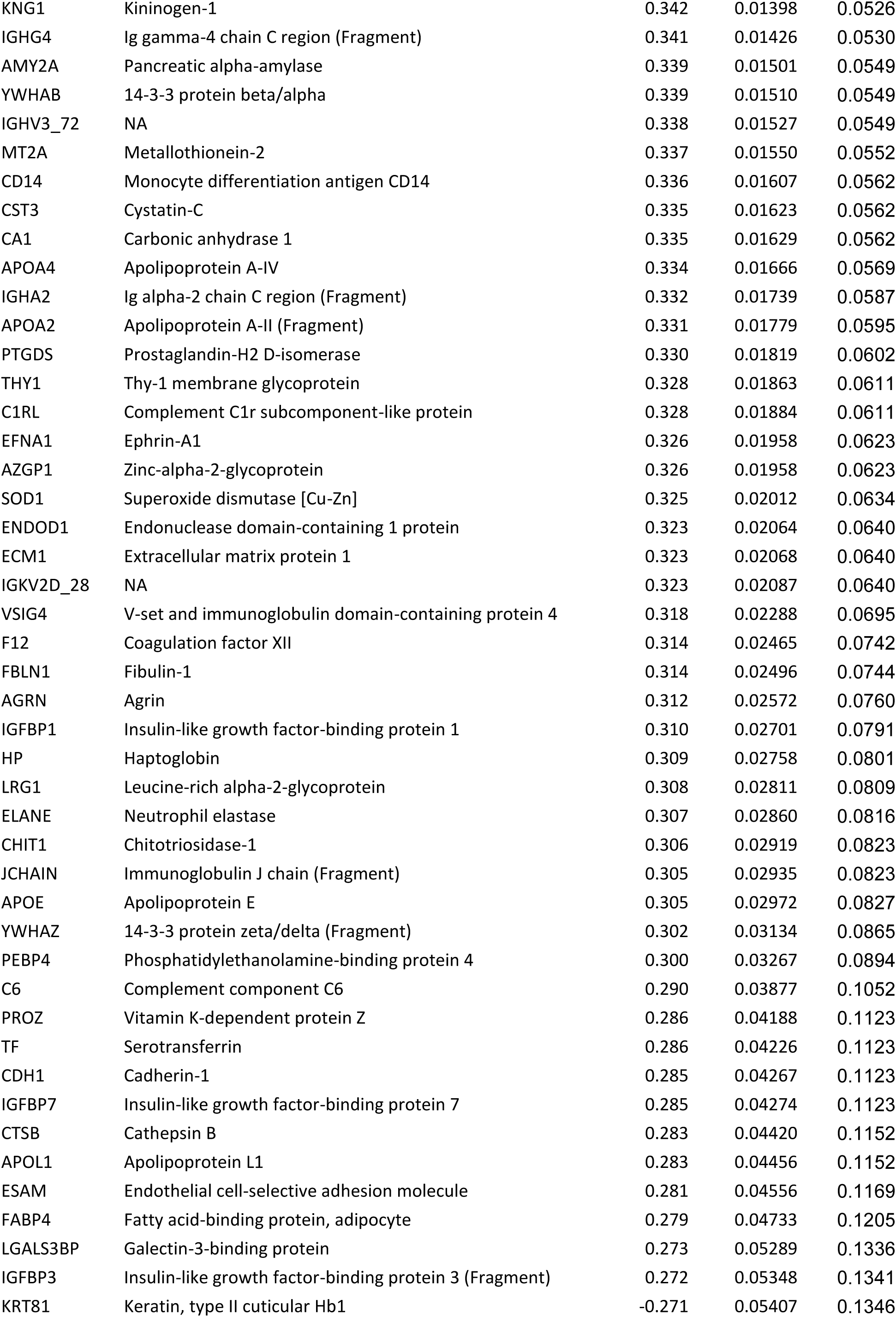
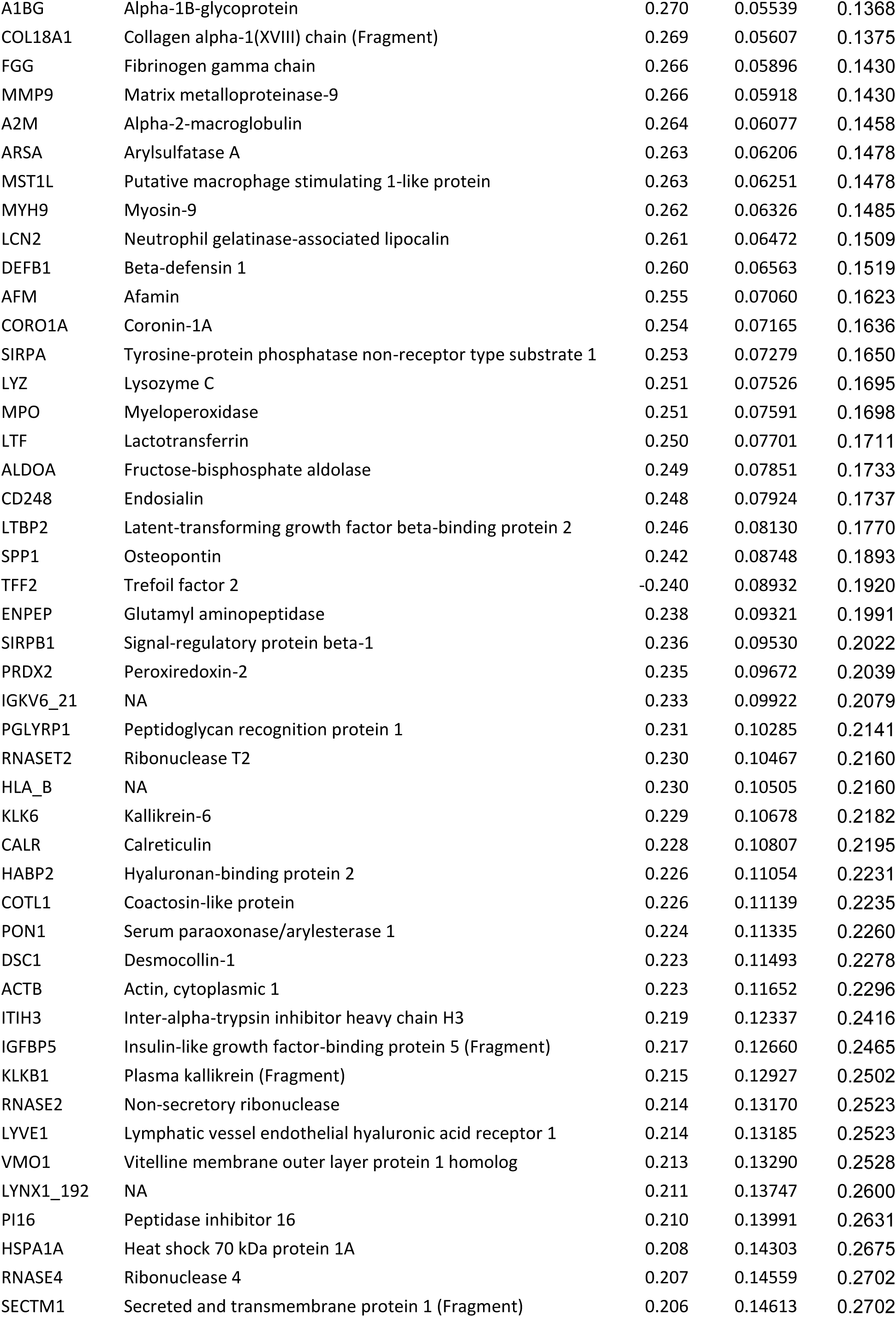
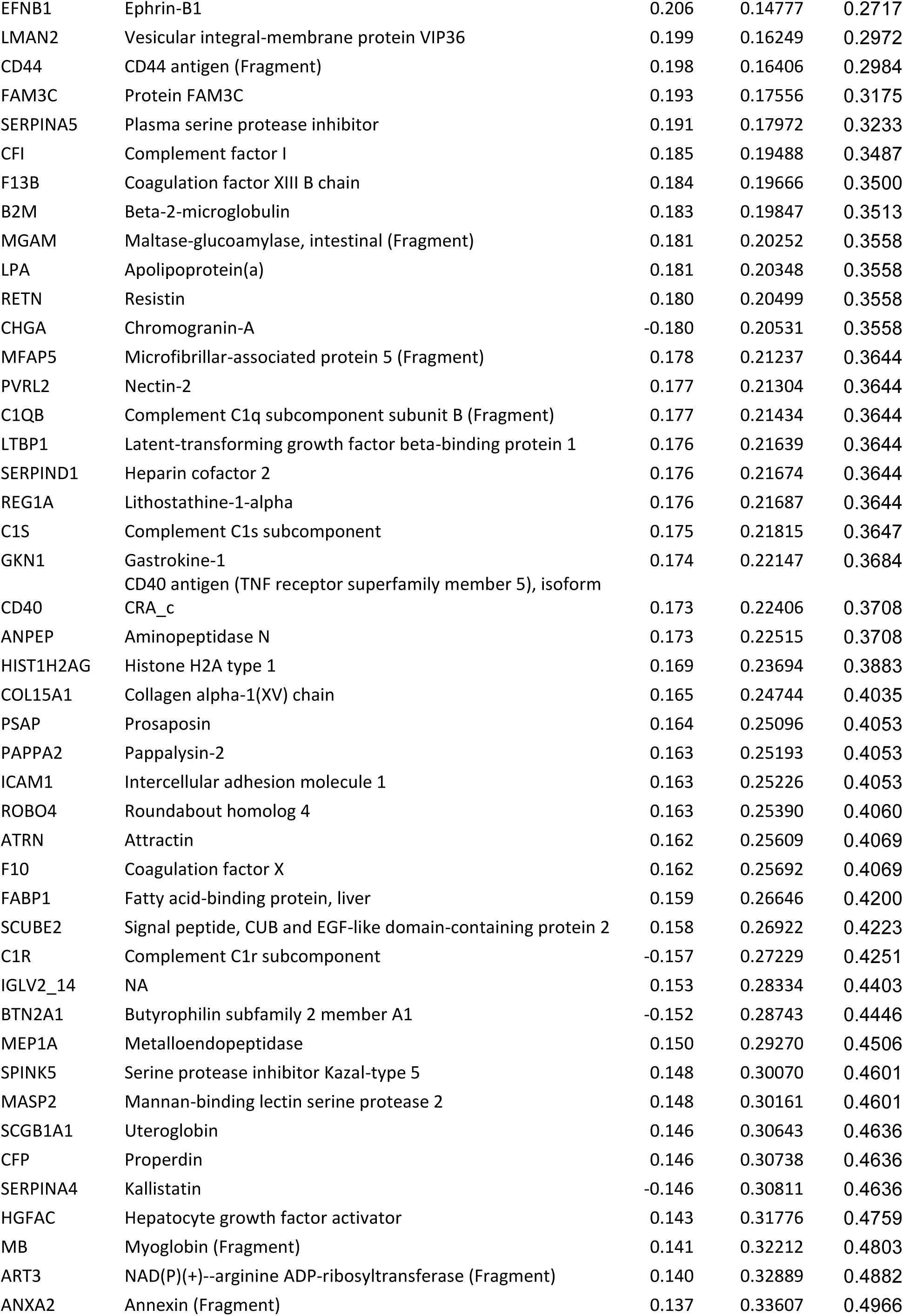
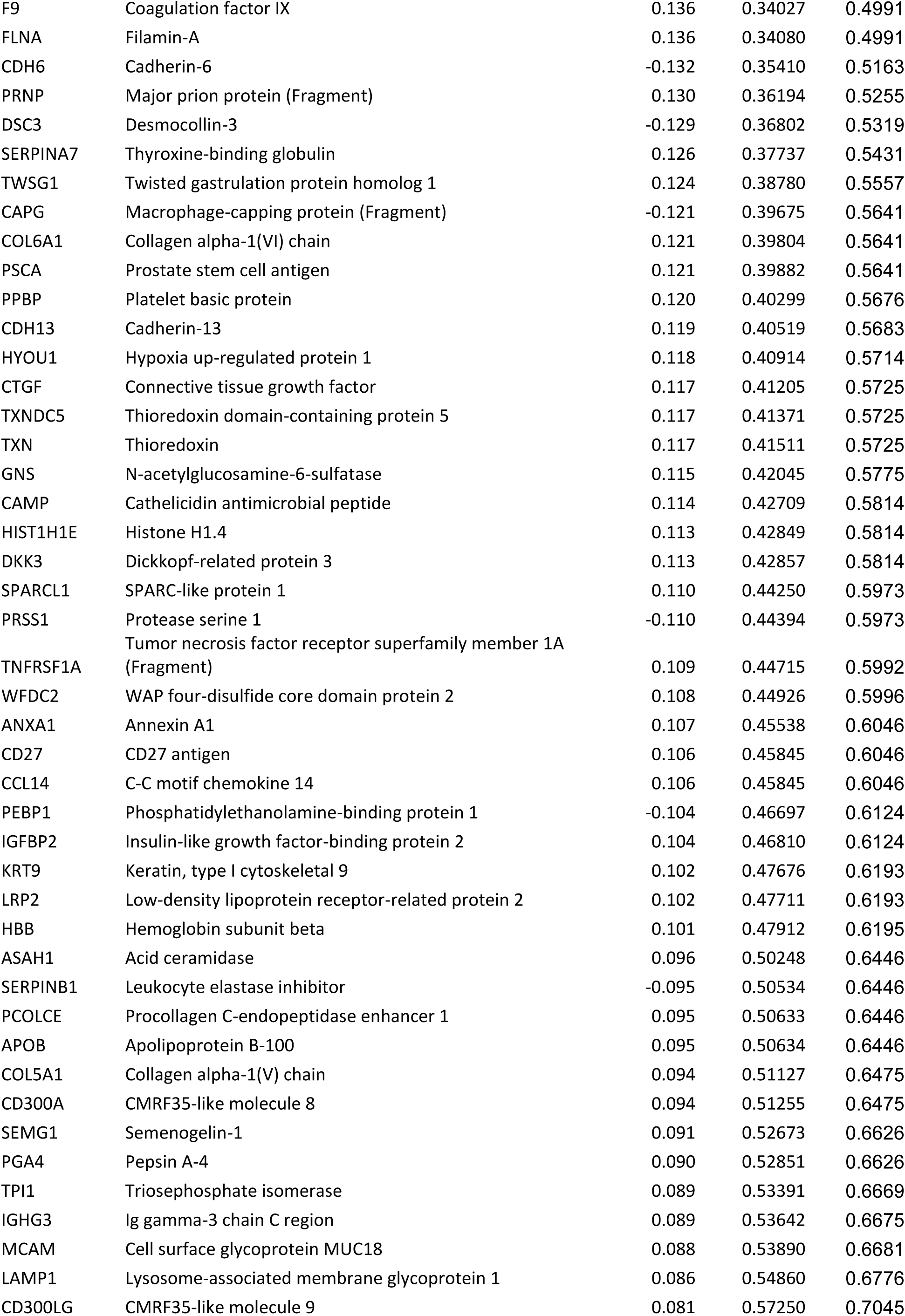
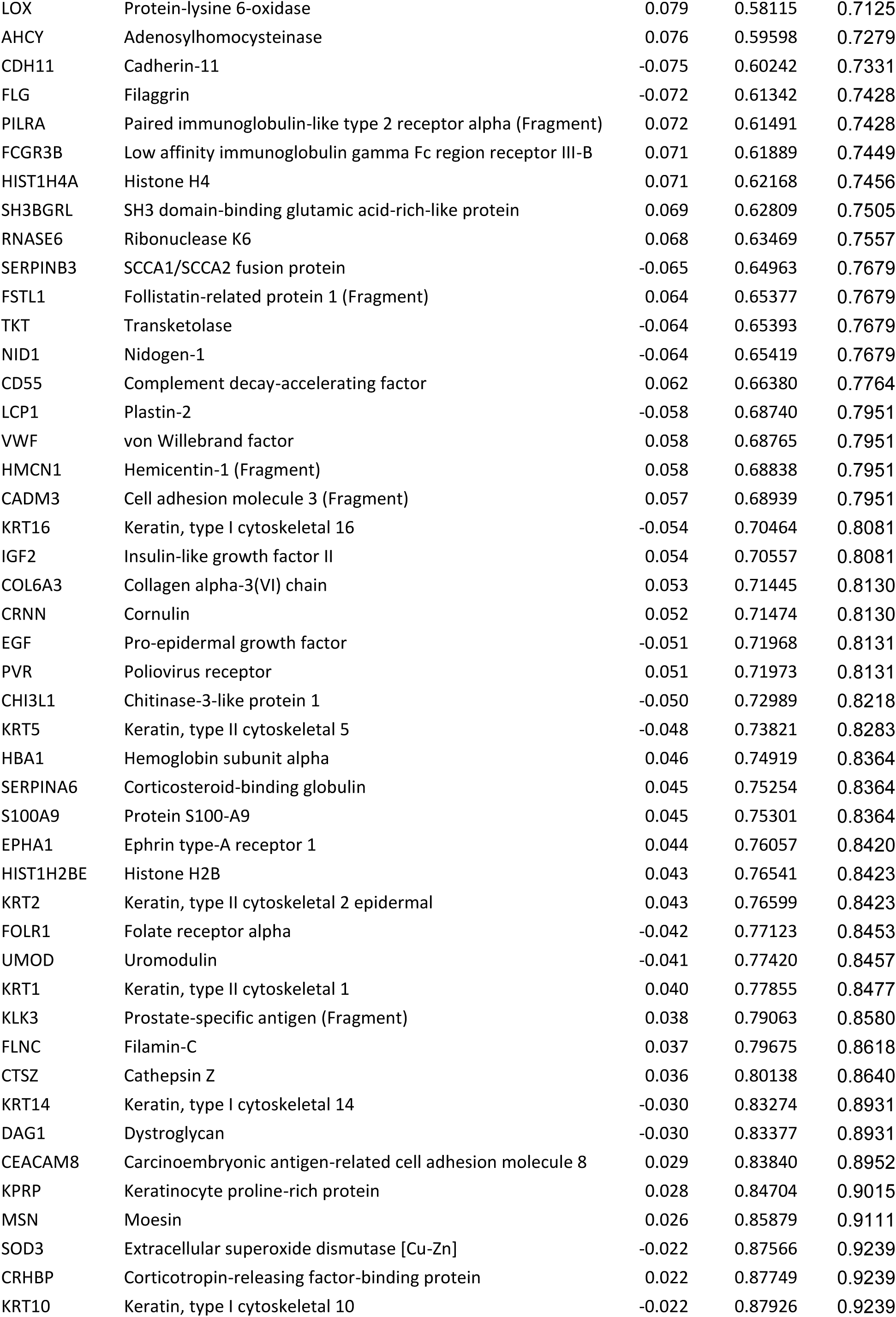
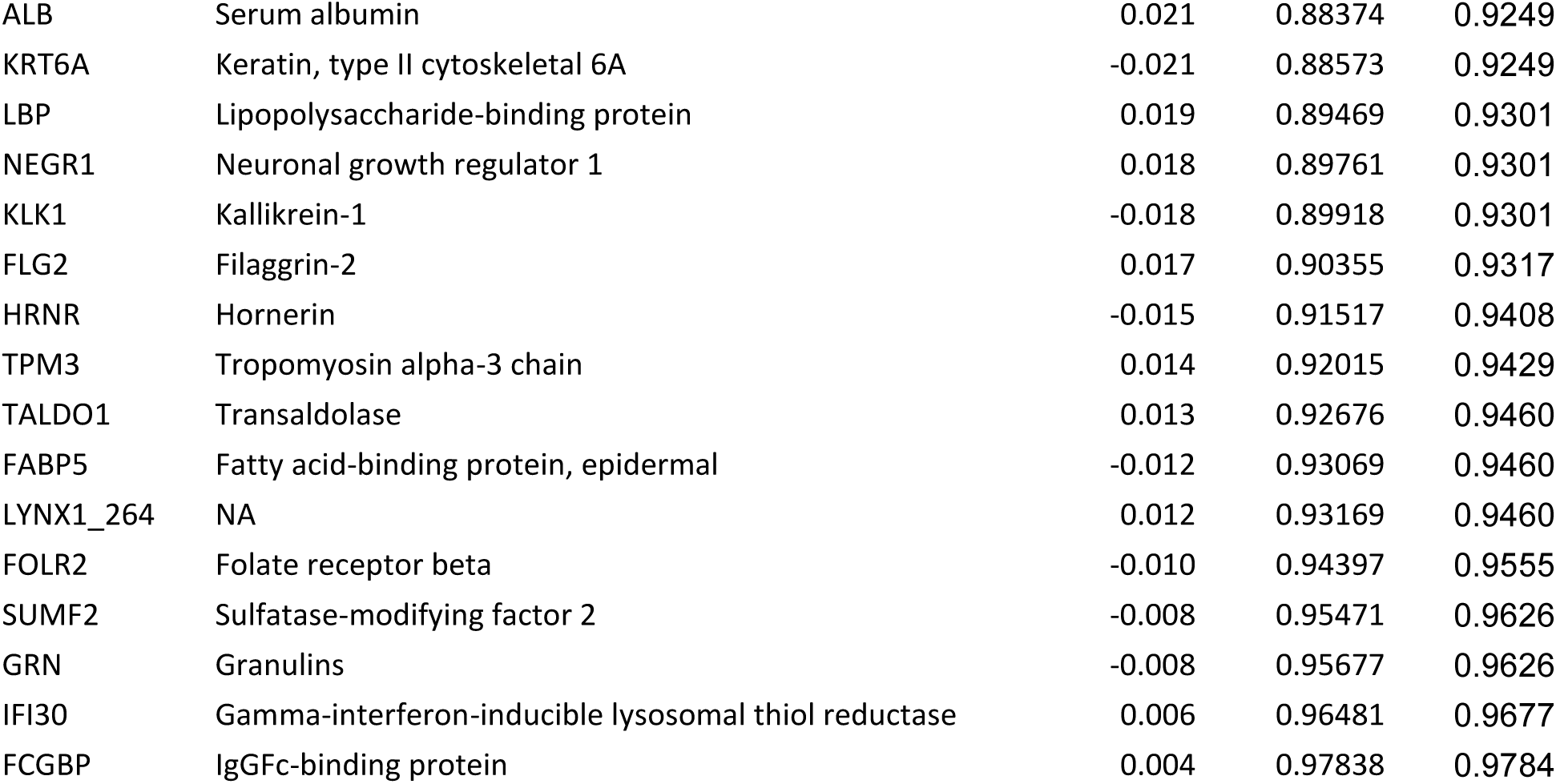
LC-MS protein correlations with proportion of p21+Ki67- senescent epithelia.

**Supplemental Table S3A.** Cox Proportional Hazards models.

| **Clusterin - using threshold** |  |  |  |  |
| --- | --- | --- | --- | --- |
|  | Hazard ratio | p | Lower CI | Upper CI |
| Clusterin threshold (above vs below) | 2.234 | 0.032 | 1.071 | 4.662 |
| Baseline eGFR (mls/min) | 0.979 | 0.004 | 0.966 | 0.994 |
| LN (Albumin Creatinine Ratio) | 1.254 | 0.043 | 1.007 | 1.562 |
| Age (years) | 0.976 | 0.034 | 0.955 | 0.998 |
| Systolic Blood Pressure | 1.018 | 0.012 | 1.004 | 1.032 |
| Sex (F vs M) | 1.022 | 0.945 | 0.552 | 1.893 |

| **Clusterin - using continuous values** |  |  |  |  |
| --- | --- | --- | --- | --- |
|  | Hazard ratio | p | Lower CI | Upper CI |
| Clusterin:Creatinine ratio per 100 µg/mmol | 1.060 | 0.002 | 1.022 | 1.099 |
| Baseline eGFR (mls/min) | 0.977 | 0.002 | 0.964 | 0.991 |
| LN (Albumin Creatinine Ratio) | 1.321 | 0.005 | 1.090 | 1.602 |
| Age (years) | 0.974 | 0.021 | 0.952 | 0.996 |
| Systolic Blood Pressure | 1.020 | 0.005 | 1.006 | 1.033 |
| Sex (F vs M) | 1.102 | 0.759 | 0.593 | 2.049 |

| **Clusterin - using log tranformed values** |  |  |  |  |
| --- | --- | --- | --- | --- |
|  | Hazard ratio | p | Lower CI | Upper CI |
| Log2 (Clusterin:Creatinine) | 1.33 | 0.01 | 1.07 | 1.64 |
| Baseline eGFR (mls/min) | 0.98 | 0.01 | 0.97 | 0.99 |
| LN (Albumin Creatinine Ratio) | 1.15 | 0.24 | 0.91 | 1.46 |
| Age (years) | 0.98 | 0.02 | 0.95 | 1.00 |
| Systolic Blood Pressure | 1.02 | 0.01 | 1.00 | 1.03 |
| Sex (F vs M) | 1.07 | 0.84 | 0.57 | 1.99 |

